# Perceptions of access to harm reduction services during the COVID-19 pandemic among people who inject drugs in Chicago

**DOI:** 10.1101/2023.10.10.23296820

**Authors:** Kathleen Kristensen, Basmattee Boodram, Wendy Avila, Juliet Pineros, Carl Latkin, Mary-Ellen Mackesy-Amiti

## Abstract

**Background:** The COVID-19 pandemic amplified the risk environment for people who inject drugs (PWID), making continued access to harm reduction services imperative. Research has shown that some harm reduction service providers were able to continue to provide services throughout the pandemic. Most of these studies, however, focused on staff perspectives, not those of PWID. Our study examines changes in perceptions of access to harm reduction services among PWID participating in a longitudinal study conducted through the University of Illinois-Chicago’s Community Outreach Intervention Project field sites during the COVID-19 pandemic.

**Methods:** Responses to a COVID-19 module added to the parent study survey that assessed the impact of COVID-19 on PWID participating in an ongoing longitudinal study were analyzed to understand how study participants’ self-reported access to harm reduction services changed throughout the pandemic. Mixed effects logistic regression was used to examine difficulty in syringe access as an outcome of COVID-19 phase.

**Results:** Most participants reported that access to syringes and naloxone remained the same as prior to the pandemic. Participants had significantly higher odds of reporting difficulty in accessing syringes earlier in the pandemic.

**Conclusions:** The lack of perceived changes in harm reduction access by PWID and the decrease in those reporting difficulty accessing syringes as the pandemic progressed suggests the efficacy of adaptations to harm reduction service provision (e.g., window and mobile service) during the pandemic. Further research is needed to understand how the COVID-19 pandemic may have impacted PWIDs’ engagement with harm reduction services.

## INTRODUCTION

The COVID-19 pandemic has disproportionately affected vulnerable populations, including people who use drugs (PWUD) (1). Compared to people without a history of substance use, PWUD are at increased risk for contracting and experiencing severe outcomes from COVID-19 (e.g., hospitalizations and mortality), particularly if they have a history of underlying health conditions including cardiovascular disease, type 2 diabetes, obesity, or cancer (1). Among PWUD, people who inject drugs (PWID) are at a heightened risk of exposure to COVID-19 for two primary reasons: complex medical comorbidities and social risk factors. Complex medical comorbidities include lung diseases such as asthma, immunodeficiencies from human immunodeficiency virus (HIV) or co-infections such as tuberculosis or viral hepatitis, and cardiovascular conditions such as hypertension. Each of these comorbidities are confirmed risk factors for developing severe COVID-19 (2,3). Along with complex medical comorbidities, vulnerability of PWID is further augmented by social factors such as homelessness, incarceration, poverty, and limited access to healthcare due to drug use stigma and discrimination (4–10). Collectively, these risk factors are compounded by economic hardships exacerbated by the COVID-19 pandemic, including food insecurity, housing insecurity, and substandard sanitation conditions (3,5,11,12). For these and other reasons, research specific to COVID-19 suggests that the pandemic escalated isolation, loss of social support, and mental health decline among PWID (5,13). Compliance with public health recommendations such as physical distancing was challenging for PWID, particularly those living in public spaces, shelters, and hostels (14). Additionally, accessibility to crucial services such as social work, counseling, HIV and hepatitis C (HCV) testing, harm reduction services, and inpatient drug treatment programs was significantly reduced (5,13).

Harm reduction services, including syringe services programs (SSPs), safe injection sites (SIFs), and naloxone distribution provide significant individual and public health benefits, including preventing deaths from overdoses and preventing transmission of bloodborne infections such as HIV and HCV among PWUD and their networks. These services reduce emergency department visits and costly healthcare services and may provide or facilitate linkage to substance use treatment (e.g., medication for opioid use disorder) (15). Moreover, these services may reduce stigma associated with drug use, affect social norms among networks, and improve access to essential resources (e.g., housing assistance, food banks, legal aid, mental health services, and employment services) (16–18). In some cases, COVID-19 mitigation strategies contributed to making SSPs and medications for opioid use disorder (MOUD) treatment more accessible by expanding mobile outreach, delivering equipment via mail, relaxing syringe exchange policies, and loosening regulations on telemedicine (19,20). Other COVID-19 mitigation strategies such as social distancing and program resource restrictions (e.g., staff, supply, financial) led to reduced access to harm reduction services, particularly SSPs, resulting in fewer opportunities for HIV testing, HIV counseling, and obtaining sterile injection supplies. Since SSPs also provide a vital location for social interaction and social support, the disruption of service modality (e.g., decreasing hours of operation, number of staff, and face-to-face interactions) has led to increased feelings of isolation among PWID (19). These findings have been identified across a range of locations, including 27 SSPs spread across the Northeast, Midwest, South, and Western regions of the United States (19).

While existing research offers valuable insights into harm reduction service access during the pandemic, these studies largely focus on the perspectives of service providers (19–21). Furthermore, the limited data reported directly by PWID are currently confined to single regions of the country, such as New York (5), and further analysis is necessary to determine if the conclusions are consistent in other regions as COVID-19 policies varied greatly by location. The present study aims to examine the impact of the COVID-19 pandemic on access to harm reduction services by exploring PWIDs’ perceptions of changes in access to these services over the course of the pandemic in metropolitan Chicago, Illinois.

## MATERIAL AND METHODS

### Study setting

We use data from an ongoing longitudinal study of PWID in metropolitan Chicago that included a COVID-19 specific module in the parent study survey. The study was conducted at Community Outreach Intervention Projects (COIP), a center within the School of Public Health at the University of Illinois at Chicago that provides harm reduction services, including SSP, naloxone, HIV and HCV counseling and testing, linkage to MOUD, and case management services to PWID and their partners in the Chicago metropolitan area (includes the city of Chicago and its surrounding suburbs that span 16 counties in northeast Illinois, southeast Wisconsin, and northwest Indiana.) The study was approved by the University of Illinois Chicago’s Institutional Review Board (Protocol #2017-0388).

### COVID-19 Ordinance

In Illinois, where the study was conducted, a stay-at-home ordinance was issued on March 20^th^, 2020, requiring non-essential workers to stay at home and non-essential businesses to close. This directive prompted community-based programs such as COIP, which provides critical treatment and harm reduction services to PWID, to modify operations by reducing hours and limiting face-to-face contact. COIP staff continually worked throughout the COVID-19 pandemic within compliance of the state and city ordinances. For example, harm reduction staff wore personal protection gear (e.g, masks, googles, shields, gloves) and interacted with clients through separation barriers to minimize interruptions to essential harm reduction services they provide to PWID in Chicago. On March 10^th^, 2020, COIP temporarily reduced public access to essential services (e.g., specialized HIV care, SSP), eliminating two of four sites and reducing hours of operation at open sites by approximately 50%. In May 2020, both COIP sites transitioned to a hybrid model of in-person and remote operations and gradually restored mobile services and full staffing at all sites by June 2021, marking a return to pre-pandemic operations. The effects of these changes on the accessibility of services for PWID in metropolitan Chicago have yet to be explored during this period, and to our knowledge, no prior study has examined PWID’s perception of access to harm reduction services during the pandemic.

### Sample and Recruitment

The present study uses data from a longitudinal COVID-19 survey initiated during the pandemic as a supplementary component to an ongoing longitudinal network and geographic study of young (aged 18-30) PWID and their injection, sexual, and social support network members in Chicago and the surrounding suburbs (22). To be eligible, the primary participant (i.e., egos) had to (i) be between ages of 18-30 years old, (ii) have injected drugs at least once in the past month, (iii) proficiently speak English, and (iv) have resided in the city of Chicago or surrounding suburbs during the past 12 months. Egos were asked to recruit up to five members of their injection networks (i.e., alters); alters were eligible for the study if they were (i) at least 18 years old, (ii) proficiently spoke English, (iii) resided in the city of Chicago or surrounding suburbs during the past 12 months, and (iv) were referred to the study by an ego. Egos and enrolled alters were followed every 6 months for up to 36 months to collect sociodemographic, network, geographic, and biologic (HIV/HCV testing) data. A more detailed description of the methods for the parent study is outlined in (22).

Participants (egos and alters) completing baseline or follow-up for the parent study were additionally asked to complete a survey on COVID-19 specific questions. These questions explored the effects of the COVID-19 pandemic on PWID, examining its implications on socioeconomic status, housing, injection drug use (IDU) behaviors and practices, and access to treatment and services.

We use baseline demographic data from the parent study as well as baseline and follow-up data from the COVID-19 survey. We used city level COVID-19 mitigation strategies as well as COIP’s service changes during the pandemic to inform a descriptive analysis of study participants’ perceptions of the accessibility of syringes and naloxone at different points in the pandemic.

### Data Collection

The COVID-19 survey was included in the parent study survey from May 2020-December 2022. Ego participants were given the opportunity to retake the COVID-19 survey at each remaining 6-month follow-up visit of the parent study, while alters participated once. Participants were recruited from the COIP field site located on the West side of Chicago. COIP provides services including syringe exchange, substance use counseling, and HCV and HIV testing and counseling as well as conducting research with PWID. The field sites are located in Chicago neighborhoods with high HIV, HCV, sexually transmitted infection incidence rates, and drug-related arrests (12). Informed consent was obtained from all study participants prior to data collection.

### Measures

The present study examines demographic measures from the baseline responses to the Alter-ego parent study survey in addition to measures from the COVID-19 sub-study of drug use behaviors, perceptions of drug use risk during the COVID-19 pandemic, and perceptions of access to harm reduction services during the COVID-19 pandemic. All responses are self-reported.

#### Demographics

Demographic characteristics were taken from the baseline parent study survey responses and included age, gender, and race/ethnicity. Race/ethnicity was categorized as non-Hispanic white, non-Hispanic Black, Hispanic, and mixed race/other. Gender was reported using three categories—male, female, transgender; for the purposes of analysis, this measure was dichotomized into the categories of male and non-male.

#### Drug Use Behaviors during COVID-19

Drug use behaviors as assessed in the COVID-19 survey were examined. Participants were asked to report if they were currently using drugs (*yes/no*) or currently injecting drugs (yes/no) Participants were also asked if their injection frequency had changed as a result of COVID-19 (*decreased a lot, decreased somewhat, decreased a little, stayed the same, increased a little, increased somewhat, increased a lot*). and if their use of new, sterile syringes had changed (*decreased a lot, decreased somewhat, decreased a little, stayed the same, increased a little, increased somewhat, increased a lot*). For analysis, injection frequency was operationalized to indicate if injection frequency increased as a result of the pandemic (*yes/no*), and sterile syringe use was operationalized to indicate if sterile syringe use increased as a result of the pandemic (*yes/no*).

#### Perceptions of Drug Use Risk during COVID-19

Two questions in the COVID-19 survey addressed participants’ perceptions of risk related to their drug use. Participants were asked if, since the onset of the COVID-19 pandemic, people are more likely to share syringes (*more likely, neither more nor less likely, less likely*). Participants were also asked if the COVID-19 pandemic has changed the degree to which they are concerned about overdose (*decreased a lot, decreased somewhat, decreased a little, stayed the same, increased a little, increased somewhat, increased a lot*). For analysis, syringe sharing was operationalized to indicate if participants thought individuals were more likely to share syringes (yes/no), and concern about overdose as a result of COVID-19 was operationalized to indicate if concern increased (*yes/no*).

#### Perceptions of Access to Harm Reduction Services

Participants’ perceptions of access to harm reduction services (e.g., sterile syringes, naloxone) as reported in the COVID-19 survey were assessed. Regarding sterile syringe access, participants were asked how their access to syringes now compares to their access prior to the onset of the COVID-19 pandemic (*access is less difficult now, access is about the same, access is more difficult now*). For analysis, syringe access was operationalized to indicate if participants perceive syringe access as more difficult than prior to the pandemic (*yes/no*). Participants were also asked if their source for obtaining new syringes has changed since the onset of the COVID-19 pandemic (*yes/no*). Regarding naloxone access, participants were asked if their access to naloxone has changed since the onset of the COVID-19 pandemic (*yes/no*). Additionally, participants were asked if they were aware of online sources of Narcan/naloxone and harm reduction supplies (*yes/no*).

### Statistical Analysis

Analyses for this study were conducted in SPSS version 28.0.1 and Stata 18.0. Prior to conducting descriptive analyses, we compared the respondents of the parent study baseline survey enrolled prior to the onset of the pandemic to those enrolled after the pandemic began to ascertain if there were any significant differences in their demographic characteristics, drug use, injection behaviors, or harm reduction usage profiles and found none.

Descriptive analysis of baseline COVID-19 survey participants (N=182) was then conducted. For categorical variables, response frequencies were calculated, and for continuous variables the mean and standard deviation were calculated. For variables that were only relevant to participants who reported engaging in IDU at the time of the survey, frequency percentages were based on the subset of individuals who responded “yes” to the question “Are you currently injecting drugs” (N=149). For variables that were only relevant to participants currently using drugs, but not specifically injection drug use, frequency percentages were based on the subset of individuals who responded “yes” to the question “Are you currently using drugs” (N=159).

Next, baseline responses across Chicago COVID-19 phases were compared using Chi-square and Fisher exact tests for all variables except age, which used independent sample t-tests to compare means. This analysis was conducted to determine if participant responses to measures of interest changed based on when during the COVID-19 pandemic their survey was conducted. Chicago COVID-19 phases were identified using the dates from press releases from the City of Chicago starting in March of 2020 and ending in March of 2022. Four major phases of the COVID-19 policy in Chicago were identified, as displayed in Figure 1. For the purposes of analysis, phases 1 and 2 and phases 3 and 4 were combined to create a pre-vaccine stage (stage 1) and a post-vaccine phase (stage 2). These phases were combined for analysis because i) phases 1 and 4 were significantly shorter periods of time than phases 2 and 3 and creating two phases resulted in two more equal time periods, and ii) there were many fewer responses to the COVID-19 survey during phase 1 (N=18) and phase 4 (N=10). All measures were then compared between the pre-vaccine (N=96) and post-vaccine (N=86) phases using Chi-square and Fisher exact tests and independent sample t-tests.

**Figure 1:**
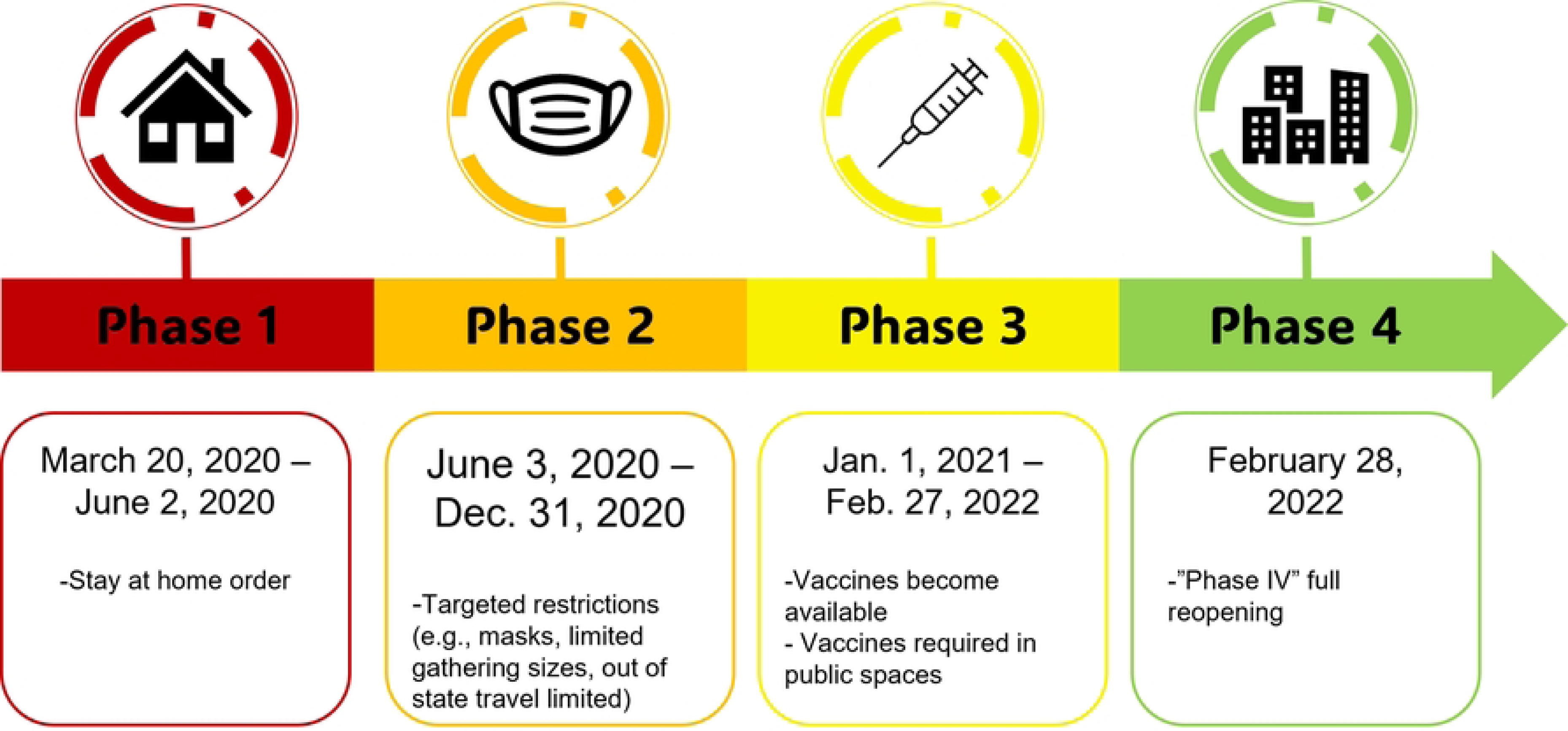
Chicago COVID-19 Mitigation Policy Timeline.

Mixed effects logistic regression with random subject intercepts was used to further examine syringe access difficulty in repeated assessments over time as predicted by COVID-19 phase. In addition to COVID-19 phase, other potential covariates were explored using Chi-square and Fisher exact tests and independent sample t-tests to compare frequencies and means by perceived syringe access difficulty for baseline responses. Cluster robust variance estimators were used to estimate standard errors and 95% confidence intervals.

## RESULTS

### Baseline Descriptive Analyses

Table 1 displays the results of the descriptive analysis of baseline responses to the COVID-19 survey as well as the results of the comparison of responses by COVID-19 phase. The sample was primarily male (70.6%) and non-Hispanic white (57.6%) with a mean age of initiation into parent study at 28.1 years old. Most of the sample reported currently injecting drugs (81.9%), with significantly (p=0.011) more participants doing so post-vaccine availability (phase 2) (89.5%) than pre-vaccine (phase 1) (75%). A sizeable proportion (32.2%) also reported increasing injection frequency to some degree, though not statistically significant.

**Table 1:**
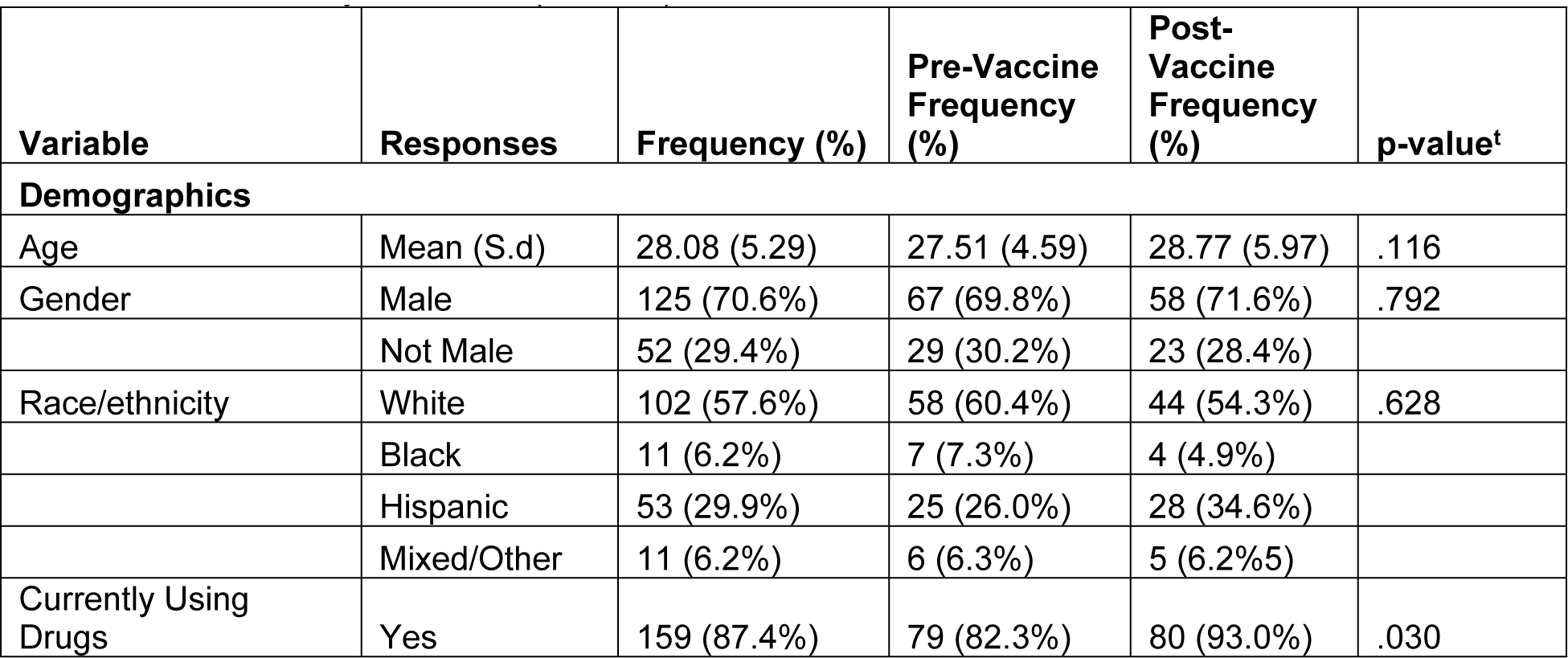
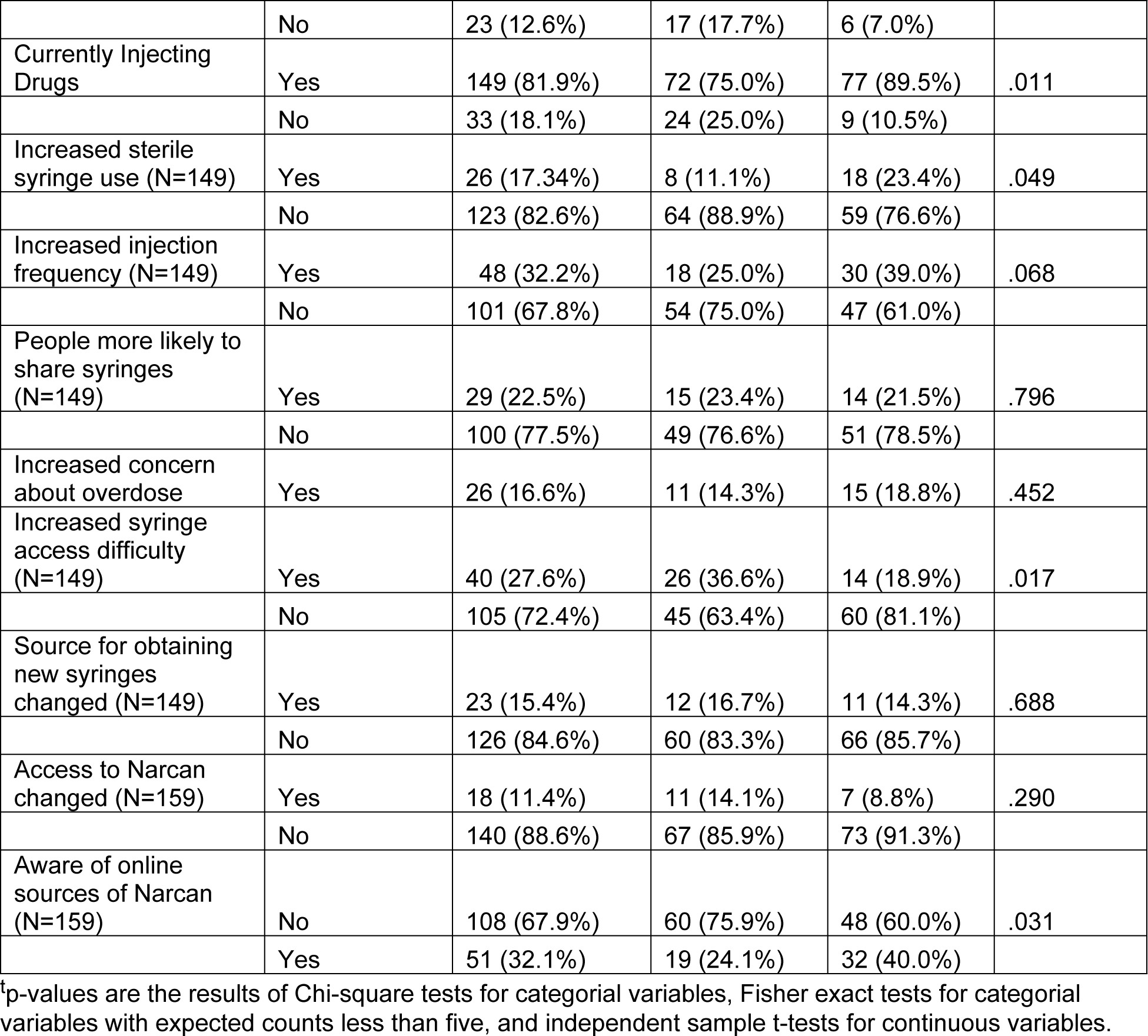
Description of Baseline Survey Results and Comparison of COVID-19 Phase 1 and Phase 2 Survey Results. (N=182)

| Variable | Responses | Frequency (%) | Pre-Vaccine Frequency (%) | Post-Vaccine Frequency (%) | p-value<sup>t</sup> |
| --- | --- | --- | --- | --- | --- |
| **Demographics** |  |  |  |  |  |
| Age | Mean (S.d) | 28.08 (5.29) | 27.51 (4.59) | 28.77 (5.97) | .116 |
| Gender | Male | 125 (70.6%) | 67 (69.8%) | 58 (71.6%) | .792 |
|  | Not Male | 52 (29.4%) | 29 (30.2%) | 23 (28.4%) |  |
| Race/ethnicity | White | 102 (57.6%) | 58 (60.4%) | 44 (54.3%) | .628 |
|  | Black | 11 (6.2%) | 7 (7.3%) | 4 (4.9%) |  |
|  | Hispanic | 53 (29.9%) | 25 (26.0%) | 28 (34.6%) |  |
|  | Mixed/Other | 11 (6.2%) | 6 (6.3%) | 5 (6.2%) |  |
| Currently Using Drugs | Yes | 159 (87.4%) | 79 (82.3%) | 80 (93.0%) | .030 |
|  | No | 23 (12.6%) | 17 (17.7%) | 6 (7.0%) |  |
| Currently Injecting Drugs | Yes | 149 (81.9%) | 72 (75.0%) | 77 (89.5%) | .011 |
|  | No | 33 (18.1%) | 24 (25.0%) | 9 (10.5%) |  |
| Increased sterile syringe use (N=149) | Yes | 26 (17.34%) | 8 (11.1%) | 18 (23.4%) | .049 |
|  | No | 123 (82.6%) | 64 (88.9%) | 59 (76.6%) |  |
| Increased injection frequency (N=149) | Yes | 48 (32.2%) | 18 (25.0%) | 30 (39.0%) | .068 |
|  | No | 101 (67.8%) | 54 (75.0%) | 47 (61.0%) |  |
| People more likely to share syringes (N=149) | Yes | 29 (22.5%) | 15 (23.4%) | 14 (21.5%) | .796 |
|  | No | 100 (77.5%) | 49 (76.6%) | 51 (78.5%) |  |
| Increased concern about overdose | Yes | 26 (16.6%) | 11 (14.3%) | 15 (18.8%) | .452 |
| Increased syringe access difficulty (N=149) | Yes | 40 (27.6%) | 26 (36.6%) | 14 (18.9%) | .017 |
|  | No | 105 (72.4%) | 45 (63.4%) | 60 (81.1%) |  |
| Source for obtaining new syringes changed (N=149) | Yes | 23 (15.4%) | 12 (16.7%) | 11 (14.3%) | .688 |
|  | No | 126 (84.6%) | 60 (83.3%) | 66 (85.7%) |  |
| Access to Narcan changed (N=159) | Yes | 18 (11.4%) | 11 (14.1%) | 7 (8.8%) | .290 |
|  | No | 140 (88.6%) | 67 (85.9%) | 73 (91.3%) |  |
| Aware of online sources of Narcan (N=159) | No | 108 (67.9%) | 60 (75.9%) | 48 (60.0%) | .031 |
|  | Yes | 51 (32.1%) | 19 (24.1%) | 32 (40.0%) |  |

<sup>a</sup>p-values are the results of Chi-square tests for categorical variables, Fisher exact tests for categorical variables with expected counts less than five, and independent sample t-tests for continuous variables.

Sterile syringe use did significantly change between phases, with more participants reporting increased sterile syringe use post-vaccine (23.4%) than pre-vaccine (11.1%). Most participants reported that they were not more likely (77.5%) to share syringes since the onset of the pandemic. Additionally, most (83.4%) participants did not report concern about overdose increasing compared to prior to the pandemic. Most participants (84.6%) reported their syringe source location(s) to be unchanged and access to syringes during the COVID-19 pandemic (72.4%) to not be more difficult. However, there was a significant difference between phase 1 and 2 (p=.017); 18% more participants reported increased difficulty in syringe access in phase 1 of the pandemic than in phase 2.

### Mixed Effects Logistic Regression

Table 2 displays the results of exploratory analysis of potential predictors of syringe access difficulty using baseline data. Syringe access difficulty was significantly different across COVID-19 phases (p=.017). Syringe access difficulty was also significantly related to perceiving people as more likely to share syringes (p<.001). Additionally, a change in the source for obtaining new syringes was significantly associated with increased syringe access difficulty (p<.001). Age, race/ethnicity, and gender were not significantly associated with increased syringe access difficulty.

**Table 2:** Baseline Comparison of Potential Predictors of Perceived Syringe Access Difficulty (N=149)

| Variable | Responses | Increased Syringe Access Difficulty |  | p-value<sup>t</sup> |
| --- | --- | --- | --- | --- |
|  |  | Yes Freq. (%) | No Freq. (%) |  |
| Post-Vaccine Phase | Yes | 14 (35.0%) | 60 (57.1%) | .017 |
|  | No | 26 (65.0%) | 45 (42.9%) |  |
| Age | Mean (S.D.) | 27.08 (2.89) | 27.63 (4.72) | .377 |
| Gender | Male | 25 (64.1%) | 72 (70.6%) | .457 |
|  | Not Male | 14 (35.9%) | 30 (29.4%) |  |
| Race/ethnicity | White | 24 (61.5%) | 63 (61.8%) | .456 |
|  | Black | 1 (2.6%) | 8 (7.8%) |  |
|  | Hispanic | 11 (28.2%) | 28 (27.5%) |  |
|  | Mixed/Other | 6 (4.3%) | 3 (2.9%) |  |
| Increased sterile syringe use | Yes | 7 (17.5%) | 17 (16.2%) | .850 |
|  | No | 33 (82.5%) | 88 (83.8%) |  |
| Increased injection frequency | Yes | 12 (30.0%) | 36 (34.3%) | .624 |
|  | No | 28 (70.0%) | 69 (65.7%) |  |
| People more likely to share syringes | Yes | 16 (43.2%) | 11 (12.5%) | <.001 |
|  | No | 21 (56.8%) | 77 (87.5%) |  |
| Source for obtaining new syringes changed | Yes | 16 (40.0%) | 7 (6.7%) | <.001 |
|  | No | 24 (60.0%) | 98 (93.3%) |  |

<sup>†</sup>p-values are the results of Chi-square tests for categorical variables, Fisher exact tests for categorical variables with expected counts less than five, and independent sample t-tests for continuous variables.

Table 3 shows the results of a mixed effects logistic regression model with random intercepts. Odds ratios and 95% confidence intervals based on cluster robust standard error estimates are reported. Individuals in the post-vaccine phase had significantly lower odds of reporting increased difficulty in accessing syringes (OR: 0.28; p=.003). Additionally, a changed source for obtaining new syringes resulted in significantly higher odds of reporting increased difficulty in access syringes (OR: 7.49; p<.0005). Perceiving people as more likely to share syringes also resulted in increased odds of reporting increased difficulty in accessing syringes (OR: 2.29; p=.040). Gender, age, and race/ethnicity did not significantly change the odds of reporting increased difficulty in access syringes.

**Table 3:** Results of Mixed Effects Logistic Regression Predicting Syringe Access.

|  | OR | 95% Confidence Interval |  | p-value |
| --- | --- | --- | --- | --- |
| Fixed Effects: |  |  |  |  |
| Post-Vaccine Phase | 0.28 | 0.12 | 0.65 | .003 |
| Source for obtaining new syringes changed | 7.49 | 2.50 | 22.48 | <.0005 |
| People more likely to share syringes | 2.29 | 1.04 | 5.05 | .040 |
| Male | 0.60 | 0.28 | 1.27 | .182 |
| Age | 0.97 | 0.90 | 1.05 | .523 |
| White | 0.53 | 0.04 | 6.85 | .624 |
| Black | 0.35 | 0.02 | 6.75 | .487 |
| Hispanic | 0.39 | 0.03 | 5.84 | .497 |
|  | Variance | Robust Std. Err. |  |  |
| Random Effects: |  |  |  |  |
| Intercept | .05 | .69 |  |  |

## DISCUSSION

Our study examined changes in perceptions of access to harm reduction during the COVID-19 pandemic among a sample of people who inject drugs in Chicago. Analyzing baseline responses, we found that fewer participants during phase 2 of the COVID-19 pandemic in Chicago indicated difficulty in accessing syringes than in phase 1. This finding was supported by mixed effects logistic regression results which showed significantly decreased odds in reporting increased difficulty in accessing syringes during the post-vaccine COVID-19 phase. Changing syringe sources and perceiving individuals to be more likely to share syringes also resulted in increased odds of reporting increased difficulty in accessing syringes. These findings indicated that, among this sample of PWID, access to syringes was perceived as less difficult as the pandemic progressed, and that individuals who reported changing syringe sources and reported that people increased syringe sharing during the pandemic were also more likely to report increased difficulty in accessing syringes.

The change in perceptions of access to syringes across COVID-19 phases can be interpreted through both changes to Chicago city policy as well as the policy changes at COIP (as described in the Introduction), which is a primary source of harm reduction services for many study participants. The stay-at-home order that was in effect from March, 2020 through the beginning of June, 2020 likely limited both study participants’ ability to access syringe exchange programs as well as programs’ ability to remain staffed and open (as evidenced by changes in COIP service provision). Existing literature shows that many syringe exchange programs in other locations were quick to adapt their programs through window, mobile, and telehealth services to minimize interruptions to service. This was the case at COIP as well, where needle exchange services were never stopped, and a window service had been established to continue to provide syringes (19,20). In addition, as the pandemic progressed, COIP increased their mobile services. These quick policy adaptations at COIP could explain why most participants did not perceive syringe access as changing during the pandemic. Additionally, the stay-at-home order lifting and staff returning to COIP as well as mobile services expanding could explain why more individuals in the beginning of the pandemic reported difficulty in accessing syringes as opposed to later in the pandemic.

The results of this study align with the extant literature on harm reduction access changes during the pandemic from the perspective of service providers. The present study’s results indicate that for most PWID in this sample, access to syringes and Narcan did not become more difficult during the pandemic supporting the efficacy of window and mobile syringe programs suggested by previous research. While it is important that individuals did not perceive significant barriers to access to harm reduction services, perceptions in access may not reflect the realities of engagement in the services. Some studies have reported increases in overdose and risk behaviors among PWID in Chicago during the pandemic, suggesting engagement in harm reduction services may be lower as a result of the pandemic despite perceptions of access not changing (5,23). Factors that influence risk taking behaviors and engagement in harm reduction such as housing stability, mental health, and social support worsened, which increased barriers to engagement in harm reduction services resulting in poorer health outcomes for PWID (5). In the case of the present study, individuals who responded to this survey were doing so as part of engagement in a harm reduction service provider (COIP), meaning that these are individuals who were able to receive these services and sought them out. While these individuals mostly reported harm reduction access as remaining the same as prior to the pandemic, they already had the ability to engage in these resources prior to seeking access. While many PWID are able to access essential harm reduction services such as clean syringes and Narcan, it is also essential to understand what barriers during the COVID-19 pandemic may have prevented PWID from engaging with these services.

Furthermore, while the results of this study indicate that adaptations to standard SSP services through window service and mobile service are effective ways of providing harm reduction services to PWID in Chicago, this study did not examine the impacts of the COVID-19 pandemic on participants’ perceptions of access to other services at COIP such as in-person counseling and HIV and HPV care. Prior studies from the perspective of harm reduction providers have shown that these in-person services were greatly reduced during COVID-19. Future research should consider how the loss of these in-person services may have impacted PWID during the pandemic and specifically how they may have impacted continued engagement in harm reduction services.

### Limitations

The current study is primarily limited by its sampling methods, which reduces the generalizability of the results to populations outside of the sample of PWID who participated in the study. Furthermore, this study recruited individuals and administered surveys through a harm reduction service provider (COIP). This biases the sample toward individuals who engaged in harm reduction services. Additionally, measures in the study reflected participants’ perceptions which means measurement error may occur due to different interpretations of, for example, what it means for something to be difficult or increase or decrease. Also, all survey questions were self-report, which creates the potential for response bias such as social desirability bias.

## CONCLUSION

The present study examined changes in PWID’s perceptions of access to harm reduction services in Chicago throughout the COVID-19 pandemic. Using data from an ongoing longitudinal study of PWID in Chicago and a COVID-19 specific sub study we analyzed participants’ perceptions of access to syringes and Narcan across phases of COVID-19 mitigation policies in Chicago as well as using mixed effects logistic regression to explore predictors of perceived increased difficulty in syringe access. The results of this analysis indicated that study participants perceived access to syringes and Narcan to be similar as compared to prior to the pandemic, while more participants found syringe access more difficult in the early part of the pandemic than later. These findings suggest that policy adaptations by COIP (the main harm reduction service provider of study participants) allowed for minimal disruption in this sample of PWID’s access to syringes and Narcan. Further research is needed to understand the effects of the pandemic on PWID’s access to other essential harm reduction services such as counseling and HIV/HPV testing as well as how the pandemic affected engagement by PWID in all harm reduction services.

## Data Availability

All relevant data are within the manuscript and its Supporting Information files.

